# The Biomedical Research Hub: A Federated Platform for Patient Research Data

**DOI:** 10.1101/2021.10.08.21264768

**Authors:** Craig Barnes, Binam Bajracharya, Matthew Cannalte, Zakir Gowani, Will Haley, Taha Kass-Hout, Kyle Hernandez, Michael Ingram, Hara Prasad Juvvala, Gina Kuffel, Plamen Martinov, J. Montgomery Maxwell, John McCann, Ankit Malhotra, Noah Metoki-Shlubsky, Chris Meyer, Andre Paredes, Jawad Qureshi, Xenia Ritter, L. Philip Schumm, Mingfei Shao, Urvi Sheth, Trevar Simmons, Alexander VanTol, Zhenyu Zhang, Robert L. Grossman

## Abstract

**Objective:** The objective was to develop and operate a cloud-based federated system for managing, analyzing and sharing patient data for research purposes, while allowing each resource sharing patient data to operate their component based upon their own governance rules. The federated system is called the Biomedical Research Hub (BRH).

**Methods:** The BRH is a cloud-based federated system built over a core set of software services called framework services. BRH framework services include authentication and authorization, services for generating and assessing FAIR data, and services for importing and exporting bulk clinical data. The BRH includes data resources providing data operated by different entities and workspaces that can access and analyze data from one or more of the data resources in the BRH.

**Results:** The BRH contains multiple data commons that in aggregate provide access to over 6 PB of research data from over 400,000 research participants.

**Discussion and conclusion:** With the growing acceptance of using public cloud computing platforms for biomedical research, and the growing use of opaque persistent digital identifiers for datasets, data objects, and other entities, there is now a foundation for systems that federate data from multiple independently operated data resources that expose FAIR APIs, each using a separate data model. Applications can be built that access data from one or more of the data resources.

## Background and significance

There are a variety of architectures and platforms that are used for patient data repositories that are designed to support research. These systems are sometimes called Research Patient Data Repositories (RPDR). It is helpful to distinguish three broad architectures for RPDR: centralized repositories, distributed repositories with a single data model, and distributed repositories with multiple data models.

### Centralized repository

The first approach is to build a centralized repository with a single data model and to curate and harmonize all patient data submitted by each separate data resource. The NCI Genomic Data Commons (GDC) [1,2] is an example of this type of centralized repository with a single data model and harmonized data.

### Distributed repository, single data model

A second approach is a distributed data warehouse with a single common data model that is separately implemented in a data warehouse containing patient data by each data resource. Since there is a common data model, queries for subject-level data can be sent to each data warehouse with the data returned and centrally analyzed. Examples of this type of system include PCORnet [3] and the HMO Research Network (HMORN) Virtual Data Warehouse (VDW) [4].

### Distributed repository, multiple data models

A third approach is a federated data warehouse with a centralized data model and local adapters at each contributing data resource that translate the central data model to the local data model. Cohort discovery at the level of counts is followed after all the required approval by queries to each data resource that return data for analysis. An example of this approach is the Shared Health Research Information Network (SHRINE) system developed using the open source Informatics for Integrating Biology and the Bedside (i2b2) platform running at each data resource [5].

With the growing acceptance of using cloud computing to support biomedical research [6], a fourth alternative is now starting to be used. With this example, the different resources are all cloud-based but expose a small set of standards-based APIs (that we call framework services below), including services for accessing metadata, accessing counts for cohort discovery, accessing data, and executing local analysis pipelines.

### Distributed repository, multiple data models, shared cloud-services

In this model, the various data resources are all cloud-based and built over a core set of cloud services. Each resource has its own data model and cloud-based applications access metadata and data, curate and harmonize data as necessary, and execute federated queries. A special case of this model is where all the data resources use a single common data model (distributed repository, single data model, shared cloud-services). There are two variants of this model. In the first, there is a single organization that provides the governance structure, including all necessary agreements, and manages and operates the federated system as a whole, ensuring interoperability. In the second, each organization separately manages and operates its own data resource and interoperability is achieved through framework services, through accepting a common framework for security, and through accepting a common principle for interoperability.

In this paper, we describe the design and implementation of a cloud-based distributed repository called the Biomedical Research Hub (BRH), with multiple data models, **in which each data repository is operated independently by a separate organization**. BRH achieves interoperability by: i) using a common set of cloud-base core software services (framework services); ii) using a common security framework (NIST SP 800-53r4); and, iii) agreeing to a principle (discussed below) that authorizes both users and environments to host sensitive data.

The BRH today contains over 6 PB of data from over 400,000 research participants. Importantly, the BRH is much more than a Research Patient Data Repository (RPDR), but rather enables a rich set of applications and cloud-based workspaces to be run over the data in the BRH. To our knowledge, the BRH is one of the first federated systems that uses these three principles to enable multiple organizations to each operate their own data repositories and still provide the ability of users to access data from two or more of the data repositories in the federated system.

## Methods

### BRH architecture

The Biomedical Research Hub is a data ecosystem in the sense that it is a loosely coupled collection of independent data resources that can be explored and analyzed with data portals, workspaces, notebooks, and other applications. All data resources and applications operate following the policies, procedures and controls of NIST SP 800-53[7]. The resources all use a small, core set of software services called framework services with open APIs. See Figure 1.

**Figure 1.**
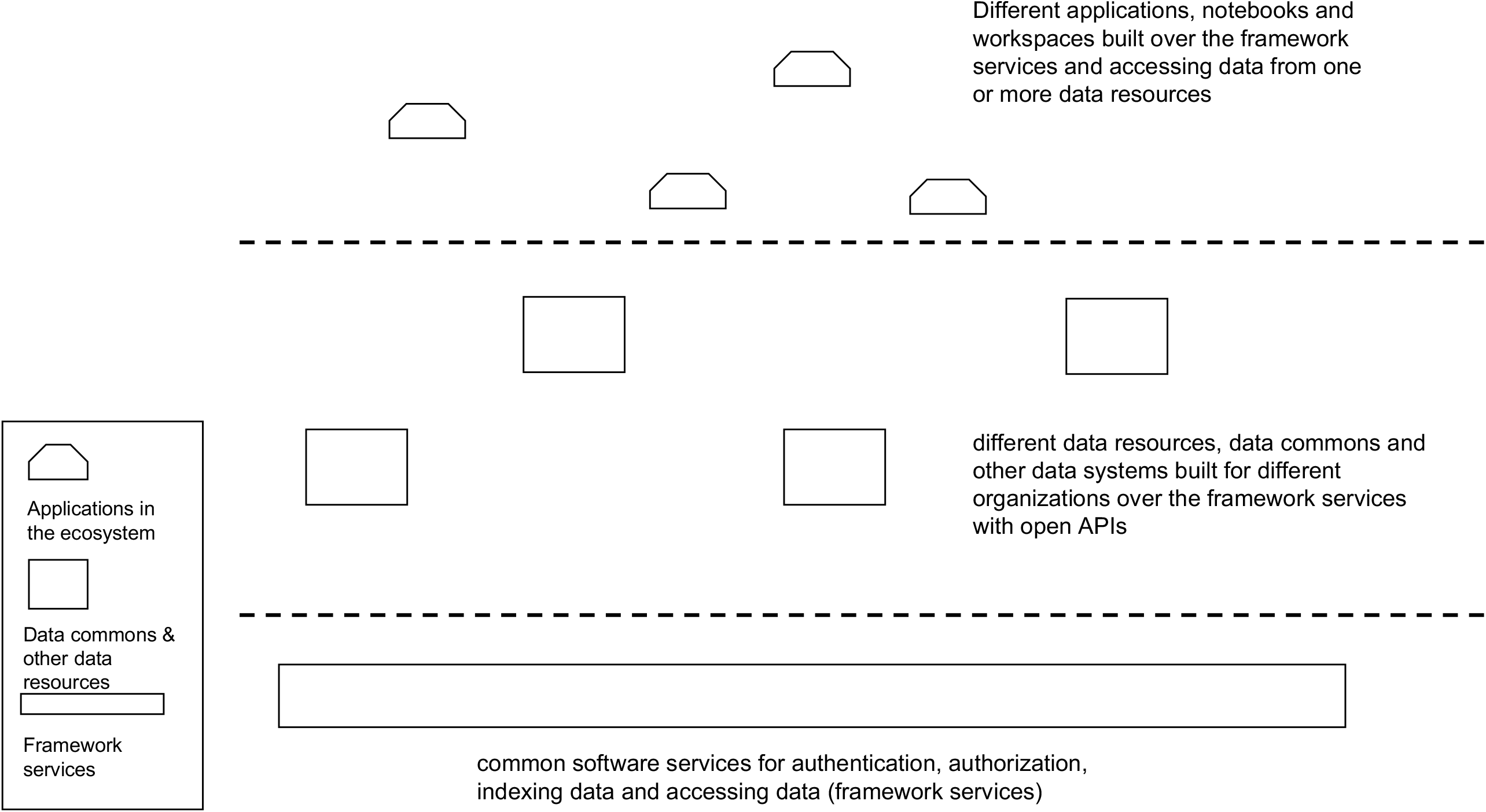
A high-level architectural overview of the cloud-based software services, data resources, and applications/workspaces in the Biomedical Research Hub.

Using the framework services and open APIs, applications can be developed for accessing, analyzing and sharing data from one or more of the data resources in the ecosystem. The framework services include services for authentication and authorization so that controlled access and other sensitive data can be accessed through the APIs.

In general, each data resource in BRH is operated by an independent organization, although some organizations operate multiple data resources. The appropriate individual(s) or committee(s) in each organization is responsible for the data governance, security, privacy, and compliance of the data resource, including managing the data contributor’s agreements, data use agreements, security and compliance decisions, etc. It is important to emphasize that each data resource and workspace operates independently on public clouds [8] with strict security and compliance policies, procedures and controls so that data from one resource are not available to a user or application associated with a second data resource unless a user is separately authorized to access a dataset in each resource and is using a workspace that is authorized and separately approved by each resource to access and manage data from that resource. This security model is sometimes called “authorize the users, authorize the environments, and trust the authorizations.” In addition, each organization separately manages its operating costs, including its cloud costs.

### Framework services

As just mentioned, the BRH was not designed as a single system, but instead as a loosely coupled collection of resources that all rely on a small set of common core software services (called framework services). There have been several attempts in the past to develop systems like these spanning NIH data resources. Our approach is broadly based on what is often called the end-to-end design principle [9,10]. This is the same approach that was used in the development of the internet and allows new sources of data and new applications that consume data to be added easily by relying on common services such as TCP, UDP, instead of adding application specific services to the system as a whole [9].

The BRH is developed using the following core framework services:

- Services for authentication
- Services for authorization
- Services for creating persistent identifiers for data and accessing data by persistent identifiers
- Services for adding, accessing and updating metadata associated with persistent identifiers.
- Services for exporting and importing bulk clinical and phenotypic data.

These services are used to connect data commons and other data resources to workspaces, which can access data from one or more data commons and other data resources. With this approach, functionality can be added to the data resources and to the workspaces without changing the overall architecture of the system.

### FAIR services for data objects

Importantly, each data resource in the Biomedical Research Hub associates a persistent digital identifier (also known as a globally unique identifier or GUID) to each dataset and each GUID is associated with metadata through a metadata service. The dataset and its metadata are accessible through an open API. In this way, data in BRH data repositories are findable, accessible, interoperable and reusable (FAIR) in the sense of [11]. We note that accessing controlled access data requires an authorization token, but is still available through an open API. In addition, some data repositories in the BRH expose a data model and support a query language interface to enable fine grained access to the data. For example, the GDC takes this approach.

Portals and other applications in the BRH are simply applications built over the framework services. For example, a portal for discovering datasets of interest in the BRH is an application over the framework services, with its primary source of information coming from the metadata service.

### Gen3 data platform

The BRH uses the open source Gen3 data platform [12], which provides the Gen3 Framework Services, Gen3 data commons for the data resources, and Gen3 Workspaces. In particular, BRH uses Gen3 Fence for authentication and authorization; Gen3 Indexd for assigning persistent identifiers to data objects and other entities and for accessing the entities; and Gen3 Metadata Services for assigning metadata to data objects and other entities with persistent identifiers.

### Data models

Each Gen3 data commons in the BRH includes a graph-based data model, with nodes and edges. Each node represents an entity, has one or more attributes representing data elements, and edges between nodes indicate relationships between them. Generally, the different data models have many common entities and data elements, but each data commons that is part of the BRH defines whatever data elements are required for the particular data they hold and for the particular applications that they support. Subject level data is available through an API via graph query language (GQL) queries [13,14]. Importantly, in general each attribute in the graph data model has a pointer to third party controlled vocabularies, such as NIH Common Data Elements [15,16].

### Sliceable data

An important feature of a Gen3 data commons is that it supports GraphQL[14] and RESTful APIs, which allow data to be queried and accessed by the slice or using range queries so that just the data needed for a particular analysis is accessed, without the requirement to import the entire dataset into a workspace.

### Governance Structure

Each data resource in the BRH sets up its own governance structure. In particular, each data resource with patient data has its own data contributor agreement (DCA) that specifies terms and conditions for contributing data to a data resource, and its own data user agreement (DUA) that specifies terms and conditions for users to access and to analyze the data. In general, the data resources in the BRH use either the NIH dbGaP agreements (NCI Genomic Data Commons, NHLBI BioData Catalyst, and the NIH Kids First Data Resource Center) or the Open Commons Consortium agreements (the VA Precision Oncology Data Commons and the Pandemic Response Commons).

An important component of the BRH governance structure is the information in an exhibit (Exhibit A) of the DCA. This exhibit, which can be customized for each dataset, specifies what type of approvals are required for accessing the data (such as whether an IRB approval is required), whether data can leave the secure boundary of the data resource and associated workspaces, what type of training is required prior to accessing the data, who can approve users to access the data (the user’s organization, the user’s PI, or the user by agreeing to the terms and conditions of the DUA), etc.

### Interoperability across BRH data resources

The ability to analyze data across multiple data resources in the BRH is achieved through several mechanisms:

1. **Common services**. Each data resource and each workspace in the BRH run a common set of Gen3 Framework Services.
2. **User authorization**. Two or more data resources within the BRH can approve a user to access their data.
3. **Environment authorization**. Two or more BRH data resources can approve a common cloud-based workspace for analyzing data.
4. **Common workflows**. Two or more BRH data resources can approve a common workflow that can be executed within a data resource’s workspace with the results returned for an integrated analysis.

### Operating Model

The BRH operating model is based on an operations center called the Commons Services Operations Center (CSOC). The CSOC is at the University of Chicago and sets up, configures, and operates each of the data resources within the BRH for each of the BRH resource sponsors using a common set of services (the Gen3 Framework Services), and a common set of security and compliance standards and SOPs. Each individual data resource is responsible for putting in place all required agreements, including data contributor agreements, data use agreements, and governance agreements, as well as for the costs to operate the data resource. Today, there are several other CSOC that operate Gen3 data commons and Gen3 workspaces and there are initial efforts to formalize how the different CSOCs can interoperate.

### Security and compliance

The University of Chicago CSOC operates all the resources in the BRH using NIST 800-53r4 policies, procedures and controls at the Moderate Level. Some of the resources in the BRH have an Authority to Operate (ATO) from their sponsors at the FISMA Moderate level, while others don’t. Regardless of whether the resources have an ATO, they operate with a common set of security and compliance requirements, which simplifies interoperability among the different resources and workspaces in the BRH.

## Results

The BRH contains a number of data resources containing patient level data at the study level, including those listed in Table 1. As already mentioned, in aggregate, the BRH contains data from over 400,000 research participants.

**Table 1.**
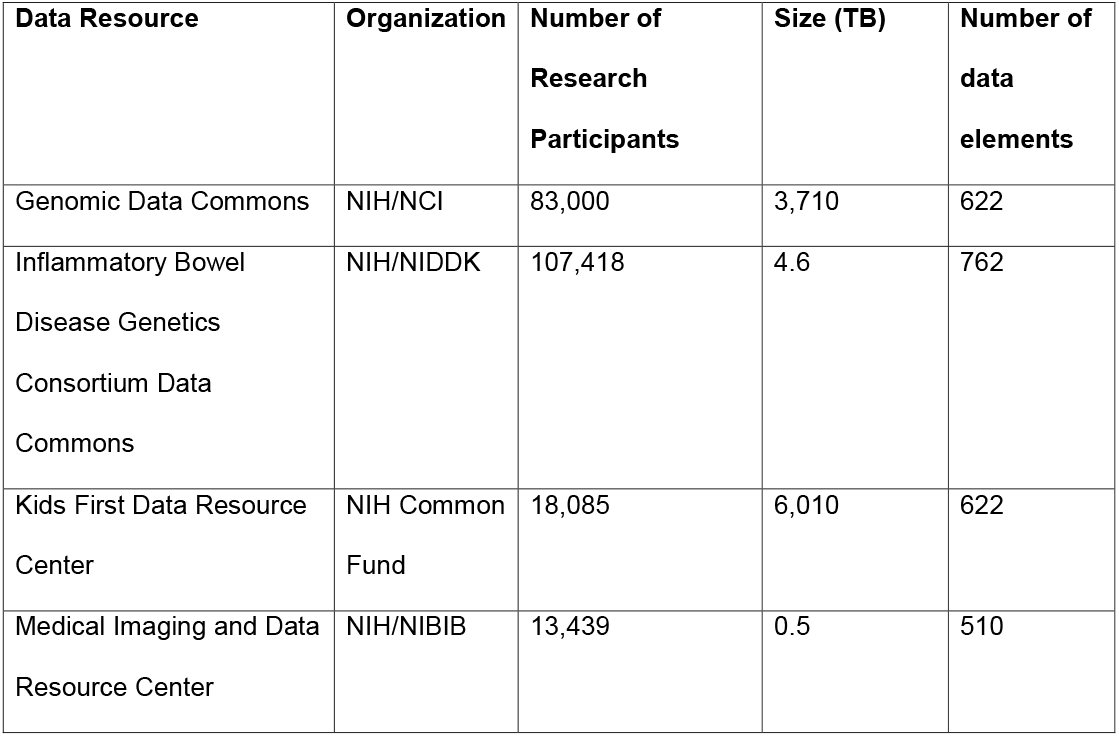
Selected data resources available in the Biomedical Research Hub.

The BRH provides a portal (the Discovery Portal) so that users can search for datasets of interest. See Figure 2. The datasets can then be loaded into a BRH workspace for analysis or downloaded. By default, BRH workspaces support Jupyter Notebooks [17], RStudio and Stata. Other tools and user-developed tools can also be added to the workspaces.

**Figure 2.**
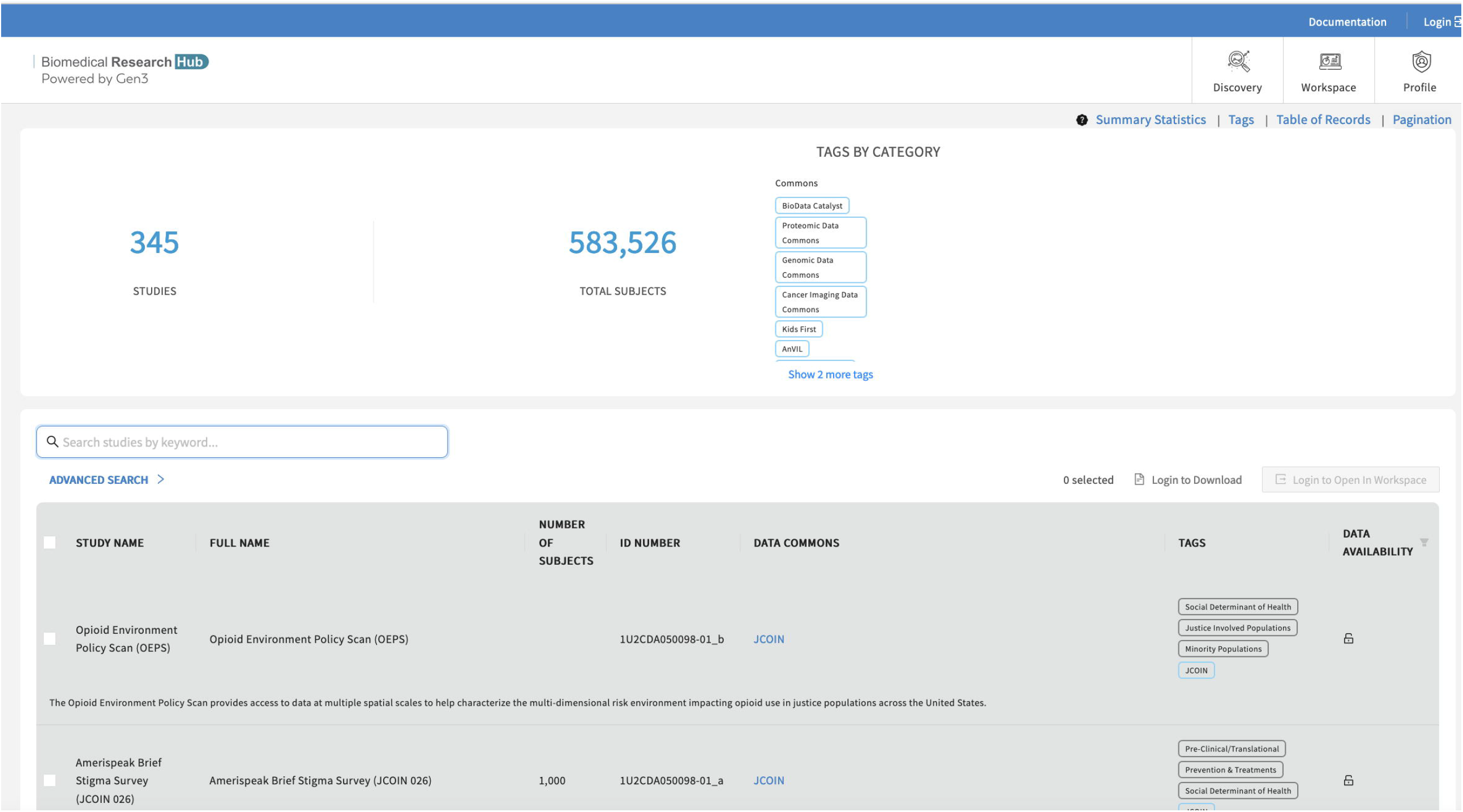
The Biomedical Research Hub Discovery Portal (https://brh.data-commons.org).

The BRH also contains data commons, which we define here following [18] as software platforms that co-locate and integrate: 1) data, 2) cloud-based computing infrastructure, and 3) commonly used software applications, tools and services to create a resource for managing, analyzing, integrating and sharing data with a community. Different data commons in the BRH contain different specialized portals and applications and interact with BRH services and applications in different ways, as the following three examples show.

### Genomic Data Commons (GDC)

As the first example, one of the data resources, the NIH National Cancer Institute (NCI) Genomic Data Commons (GDC) contains data on over 83,000 research participants and provides interactive portals for exploring and visualizing harmonized cancer genomic, clinical and imaging data about them [1]. The datasets (corresponding to projects in the GDC terminology) are all harmonized to a common GDC data model [1]. The GDC uses the NIH eRA Commons system for authentication and the NIH dbGaP [19] for authorization.The BRH interacts with the GDC in two fundamental ways. First, the GDC provides interactive tools to create virtual cohorts that can be accessed and explored from BRH workspaces using the GDC APIs. The BRH interoperates with NIH dbGaP and NIH RAS service so that researchers using BRH workspaces can be authenticated and authorized using these standard NIH services. Second, metadata about GDC datasets (i.e. GDC projects) have been added to the BRH metadata service, allowing researchers to find GDC projects of interest through the BRH Discovery Portal. These datasets can then be accessed and analyzed using BRH workspaces as just described. The GDC is built using some Gen3 software but is not a Gen3 data commons, but since it exposes an API that can be used to access both open access and controlled access data, it was straightforward for BRH to access the required metadata about GDC datasets for the BRH Discovery Portal and to access the required data from BRH workspaces.

### IBDGC Commons

As the second example, another of the BRH data resources, the NIH National Institute of Diabetes & Digestive & Kidney Diseases (NIDDK) Inflammatory Bowel Disease Genetics Consortium Data Commons (IBDGC Commons) contains harmonized data on over 110,000 research participants and provides both portals for exploring the data in the commons and workspaces for analyzing those data using standard tools. Interactive Jupyter notebooks are provided to facilitate analyses such as using single cell data to explore differences between inflamed and non-inflamed ileum in Crohn’s disease. The IBDGC Commons uses the Gen3 core FAIR data services that support the persistent identifiers, metadata, and DRS-compliant access services so that the BRH Discovery Portal and BRH workspaces can be used with data from the IBDGC Commons. In addition, the Commons has a Gen3 harmonized data model and associated GQL API so that subject level data from the IBDGC can be analyzed using BRH workspaces.

### Medical Imaging and Data Resource Center

The Medical Imaging and Data Resource Center (MIDRC) is a Gen3 commons containing imaging and associated clinical data. During its first year, MIDRC is focused on COVID-related images. Currently, MIDRC contains over 13,000 imaging studies, with about 49,000 additional imaging studies currently undergoing data quality and harmonization, and expected to be available in 2021. MIDRC uses both Gen3 FAIR Data Services as well as a harmonized Gen3 graph-based data model so that both dataset-level and subject-level data is available to BRH resources, such as BRH workspaces. MIDRC uses Gen3 services for authentication and authorization.

## Discussion

The advantage of a loosely coupled RPDR is that individual resources can develop systems that are designed to satisfy their own goals and objectives but are part of a broader ecosystem. The disadvantage is that it can be challenging to analyze data across resources since data harmonization is usually required. Table 2 summarizes some of the similarities and differences for distributed systems that manage research participant data.

**Table 2.** Some of the similarities and differences between the BRH and other distributed systems for managing research participant data.

|  | **Centralized or distributed data** | **Number of system operators** | **Data harmonization level** | **Data access level** |
| --- | --- | --- | --- | --- |
| Biomedical Research Hub (BRH) | Data distributed across multiple systems (one per organization) | Multiple system operators, with systems managed by separate organizations | Multiple data models, with data elements linking to Common Data Elements and controlled vocabularies | Dataset level |
| Several commercial systems | Data distributed across multiple systems (one per organization) | One system operator | One data model, with local adapters | Research participant level |
| HMO Research Network (HMORN) Virtual Data Warehouse | Data distributed across multiple systems (one per organization) | Multiple system operators with shared governance model | Single data model | Research participant level |
| NCI Genomic Data Commons (GDC) | Centralized data in one system | One system operator | Harmonized data with single data model | Research participant level |
| NCI Cancer Research Data Commons (CRDC) | Data is distributed across multiple systems (one per data type) | Multiple system operators managed by a single organization | Multiple data models (one per system), with harmonized data model (across systems) | At dataset and research participant level |

The sponsor of each data resource in the BRH can customize their resource as desired, as long as the resource uses the BRH framework services, as long as the resource follows FISMA 800-53 policies, procedures and controls for security and compliance, and as long the data resource has a process for authorizing users to access data and for authorizing environments for the analysis of data. In particular, each resource can develop their own data model, use their own data governance structure, use their own common’s governance structure, develop custom front ends and applications, etc. In other words, from the sponsor’s perspective, they have complete control over their resource, yet there is enough structure through the framework services, shared security and compliance policies, procedures and controls, and approving cloud platforms as authorized environments for the analysis of their data.

Of course, the more the data models differ between BRH data resources, the more work is required by researchers to harmonize the data from two or more resources, when this is required. In general, each data element in a Gen3 data model contains a pointer to a corresponding Common Data Element (CDE)[15], controlled vocabulary, or third party standard, such as CDISC[20]. With the use of CDEs, third party standards and controlled vocabularies, harmonizing two data models is much easier and can leverage API-based semantic services for this purpose, such as the NCI Thesaurus[16].

Sometimes, it can be useful to combine datasets from different BRH data resources. For example, a researcher seeking to understand comorbidities between IBD and cardiovascular disease [21] may want to do a combined analysis of datasets from the IBD Genetics Consortium Data Commons and selected datasets from BioData Catalyst.

BRH workspaces are designed so that they can leverage all the various native AWS services for computing, data management, machine learning, etc. Initially, some effort is required from a security and compliance perspective to bring a new service into the BRH, but after this initial effort the service is available throughout the BRH.

We have learned a number of lessons from our experience developing the individual commons, the core services that they rely on, and the BRH applications and workspaces,

1. **Federate using common core services**. Provide flexibility and autonomy for each data resource to develop their own system. Let each resource develop and customize the system that best suits their purpose, their data and systems governance, and the applications that they need, but require each system to use a core set of software services (framework services) to simplify federation and interoperability. In general, the project sponsors do not have strong opinions about the underlying frameworks services and are willing to expose APIs. In this way, each resource can be part of a federated system, but still have full control in the design and development of their resource.
2. **Support different data models, but require common data elements (with links)**. Although each resource in the BRH separately and individually develops its own data model, these are based on several community developed data models (in particular the GDCs) that provide a foundation for data models for the other resources. The data elements in the individual data models each point to third party definitions and controlled vocabularies, such as Common Data Elements (CDE), and in this way, there is a fair amount of interoperability that is provided through the common definitions, which can be supplemented by cross walking definitions through semantic services, such as the NCI Thesaurus [16,22].
3. **Expose datasets through FAIR APIs**. Each Gen3 commons provides metadata about its datasets through a metadata service available through an open API so that datasets can be easily discovered by applications such as the BRH Discovery Portal. In addition, BRH workspaces that support applications, such as Jupyter Notebooks, RStudio, and Stata, can assess, explore and analyze the datasets themselves via persistent identifiers through open APIs.
4. **Expose subject level data through APIs**. All the Gen3 commons expose subject level data through APIs, as well as the corresponding data models. With this capability and by using the shared framework services, applications to discover, explore and analyze subject level data can be developed, as has been done in the BRH. Occasionally a one time extract-transform-load may still be required to harmonize data that don’t have data elements that are shared.
5. **Resources should approve multiple third party cloud-based workspaces**. Today, there is often hesitancy to approve the analysis of data in third party cloud-based workspaces, which is one of the barriers to the wider sharing and utilization of patient research data. Data resources should evaluate the security, compliance, functionality and ease of use of different cloud based workspaces and approve several as authorized environments so that the data they host can be explored and analyzed by researchers in convenient and secure environments.

## Conclusion

We have described the design and operating model for the Biomedical Research Hub (BRH), a distributed cloud based system that integrates independent repositories for patient level data to support research with workspaces that can access data from one or more repositories. The BRH is built over a core set of common software services following the end-to-end design principle in distributed architectures. With this design, the BRH supports interoperability but also allows for new data resources and new applications to be added to the BRH without changing the core architecture.

Today, the BRH consists of multiple data commons each providing: 1) data portals for data exploration and cohort discovery and 2) workspaces for analyzing data using Jupyter notebooks and other applications. Since each of the commons expose APIs that provide FAIR discovery, access to datasets, FAIR cohort discovery and access to subject level data, the BRH also supports applications and workspaces across two or more commons. Currently, the BRH contains a BRH Discovery Portal that can search across all the commons and BRH workspaces that can access and explore data from one, two or more commons. To summarize, the BRH is an example of a Research Patient Data Repository that is cloud-based, distributed, based on a small core set of shared cloud-services, and supports multiple data models.

## Data Availability

This project did not generate data nor analyze data but developed a software platform for searching and analyzing data that is available through data resources that expose APIs for discovering and accessing data.

## Funding Statement

Research reported in this publication was supported in part by the NIH Common Fund under Award Number U2CHL138346, which is administered by the National Heart, Lung, And Blood Institute of the National Institutes of Health. The content is solely the responsibility of the authors and does not necessarily represent the official views of the National Institutes of Health.

The Research was also funded in part by the Amazon Diagnostic Development Initiative (DDI) in an award to the Open Commons Consortium, a division of the Center for Computational Science Research, Inc. (CCSR).

## Competing Interests Statement

The authors have no competing interests.

## Contributorship Statement

All the authors contributed to the conceptualization, implementation or description of the Biomedical Research Hub system.

## Notes

### Competing Interest Statement

The authors have declared no competing interest.

